# Evaluating the buffering role of perceived social support and coping resources against the adult mental health impacts of COVID-19 psychosocial stress: a cross-sectional study in South Africa

**DOI:** 10.1101/2023.06.20.23291688

**Authors:** Andrew Wooyoung Kim, Someleze Swana, Mallika S. Sarma

## Abstract

**Objectivesw:** Growing evidence has highlighted the global mental health impacts of the COVID- 19 pandemic and lockdown, particularly in societies with pre-existing socioeconomic adversities and public health concerns. Despite the sudden and prolonged nature of many psychosocial stressors during the pandemic, recent studies have shown that communities utilized several coping mechanisms to buffer the mental health consequences of COVID-related stress. This paper examines the extent to which coping resources and social support buffered against the mental health effects of COVID-19 psychosocial stress among adults in South Africa.

**Materials & Methods:** Adult participants (n=117) completed an online survey during the second and third waves of the COVID-19 pandemic in South Africa (January-July 2021), which assessed experiences of stress, coping resources, social support, and four mental health outcomes: depression, anxiety, post-traumatic stress disorder, and bipolar disorder. Moderation analyses examined the potential buffering role of coping resources and social support against the mental health effects of COVID-19 stress.

**Results:** Adults reported elevated rates of psychiatric symptoms. Coping resources buffered against the poor mental health effects of COVID-19 psychosocial stress, whereas perceived social support did not significantly moderate the association between COVID-19 stress and adult mental health.

**Discussion:** These results suggest that adults in our sample utilized a variety of coping resources to protect their mental health against psychosocial stress experienced during the COVID-19 lockdown and pandemic in South Africa. Additionally, existing mental health conditions and strained social relationships may have attenuated the potential stress-buffering effect of perceived social support on adult mental health.

## INTRODUCTION

The SARS-COVID-19 pandemic and subsequent lockdown severely disrupted everyday life and infrastructure across the world while introducing a wide variety of global-level stressors. Building on everyday stressors, many of the additional pressures COVID-19 brought were novel and abrupt, compounding negative effects. They were also broad reaching and deeply felt across sectors, impacting economic, social, interpersonal, and healthcare domains. Since its onset, COVID-19 has led to at least 6.55 million global deaths but also has caused upheaval through lost jobs, depletion of resources, and the cause of short-and long-term disability for an unquantified number of people. In particular, the ongoing COVID-19 pandemic has been disproportionately harmful to people at the margins already living in precarity. Several studies have demonstrated that individuals with pre-existing conditions were more likely to experience COVID-related hospitalization, ongoing morbidity, or mortality (Fang et al., 2020; Sanyaolu et al., 2020; Wang et al., 2020). Marginalized populations, particularly those that have experienced systemic and pervasive violence and trauma, have higher rates of morbidity and mortality, particularly during the COVID-19 pandemic (Brakefield et al., 2022; Braveman & Gottleib, 2014). In lower-to-middle income countries (LMIC), like South Africa, the pandemic has introduced further mental health threats compounding existing ones (Kim et al., 2022).

As of October 2022, 4.02 million cases of COVID-19 have been reported in South Africa, resulting in 102,000 deaths (Our World in Data, 2022). Like in many other parts of the world, in South Africa, the onset of the pandemic brought about violent and abrupt disruptions to economic and social resources, including travel, and gathering restrictions, as well as strain on an already burdened healthcare infrastructure. Further, as a highly contagious airborne disease, the containment and treatment of COVID-19 necessitated mandated social isolation, which has well-documented negative impacts on mental health. In South Africa, a country recovering from the violent legacies of apartheid and the subsequent downstream high rates of stress-related health conditions (Coovadia et al., 2009; Kim et al., 2023), the additional stressors stemming from COVID-19 created compounded stressors on an already overburdened healthcare system. This struggling healthcare system, and the epidemic of non-COVID-related communicable and non-communicable diseases, is an outcome of policies derived from colonial subjugation, apartheid dispossession, and post-apartheid recovery. Across these periods, racial and gender discrimination, the migrant labor system, the destruction of family life, vast income inequalities, and extreme violence have shaped health and health services (Coovadia et al., 2009; Barbarin & Richter, 2013; Kaminer & Eagle, 2010). These conditions have primed a population facing inequality across multiple sectors, including in healthcare and disease incidence, to be disproportionately impacted by COVID onset and recovery.

For many individuals, increased psychosocial stress has been a hallmark of this pandemic. The pervasive and powerful impacts of stress on different aspects of human functioning and well-being, particularly on mental health, have been well described in the literature (Lupien et al., 2009; Lupien et al., 2018). Chronic psychosocial stress has powerful effects on individual physiology including changes to sleep, metabolism, and immune function (Russel & Lightman, 2019; Sanford et al., 2023). The ubiquitous stress of the pandemic has had substantial downstream impacts on mental health in populations around the world (Hossain et al., 2020; Manchia et al., 2022; Oyenubi et al., 2022; Subramaney et al., 2020). Chronic psychosocial stress has also long been associated with negative mental health, including increased incidence of depression, anxiety, burnout, pathological aging, and post-traumatic stress disorders (Burke et al., 2005; Marin et al., 2011; McEwen, 2017; Meewisse et al., 2007; Metzger et al., 2008; Steudte et al., 2013; Uchino, 2006; Yehuda et al., 2005). These effects of psychosocial stress on mental health tend to be context-specific, where outcomes differ on an individual basis and are shaped by factors such as severity, duration, or unpredictability of the stressor. Further, untreated mental health issues can leave individuals more susceptible to future mental health problems in times of crisis, creating compounded effects.

In global crises, the context in which individual stress response is often shaped by their social world, and much of the COVID-19 experience has been characterized by social isolation. It is well known that social isolation can have devastating effects on highly social animals like humans – social isolation has been shown to transform stress response mechanisms and in the last 25 years, has been recognized as a major risk factor for morbidity and mortality in humans (Cacioppo et al., 2003; Cacioppo et al., 2015; Hostinar et al., 2015). Coping plays an important role in processing negative experiences, particularly those related to social isolation, and can shape physical and mental health (Cacioppo et al., 2003). The COVID-19 pandemic, like many other global crises, led to an increased reliance on a variety of coping mechanisms (Bhattacharjee & Ghosh, 2022; Polizzi et al., 2020). These coping mechanisms have included support from family and friends, changes in attitude (i.e., a positive outlook or acceptance), activities (i.e., staying occupied/busy, activities promoting relaxation, exercise), taking medications, religious practices, counseling/therapy, or crying.

Social support, specifically, is a powerful and well-documented coping mechanism. In settings of extreme stress, highly resilient individuals are particularly adept at forming supportive social attachments (Charney, 2004). Socially supportive ties play two major roles in times of stress: (1) helping individuals process and control emotional responses to stressful situations and (2) keeping physiological, neuroendocrine, and immunologic responses to stress at lower levels and/or by promoting faster recovery of these systems when responding to a stressor (Cohen & Wills, 1985; Taylor, 2011). Early work by Uchino and colleagues demonstrated the positive physiological effects of perceived social support/connectedness, including lower resting blood pressure, better immunosurveillance, and lower levels of basal catecholamines (Uchino et al., 1996; Hennessy et al., 2009; Taylor, 2011). Additionally, many mechanisms of social support, including informational support (i.e., helping another to understand a stressful event and available resources better), instrumental support (i.e., provisioning of tangible assistance, such as services or financial assistance), and emotional support (i.e., providing warmth and nurturance), likely ameliorate the adverse consequences of stress and trauma (Cohen & Wills, 1985; Taylor, 2011).

During disaster scenarios, particularly pandemics (e.g., HIV/AIDS, H1N1 influenza, SARS, and Ebola), coping mechanisms are key. For example, increased social support has been associated with lower rates of mental health problems in these settings (Asante, 2012; Chew et al., 2020; Guilaran et al., 2018). This is also true for COVID-19. In a study with 405 students at an American university, Szkody and colleagues report that when accounting for time in social isolation, perceived social support buffered the association between concerns about COVID-19 and psychological health (Szkody et al., 2021). In South Africa, recent work suggests that individuals embedded within care networks tended to weather the pandemic better – Steigler and Bouchard showed that those confined with family members tended to be more optimistic than those confined alone and were able to spend any leisure time doing family activities, thus staving off boredom, anxieties, and rumination on the situation (Stiegler & Bouchard, 2020).

Here, we examined the relationship between COVID-19 stress and self-reported mental health outcomes and the potential buffering effects of coping and social support in a cohort of adults living through the COVID-19 pandemic across South Africa. We were interested in testing if COVID psychosocial stress was associated with adult mental health, particularly the incidence of symptoms for four main psychiatric conditions: depression, anxiety, bipolar disorder, and post-traumatic stress disorder (PTSD). Further, using multivariate regression analyses, we evaluated if coping and social support attenuated the relationships between COVID psychosocial stress and adult mental health outcomes.

## METHODS

### Study sample

This study was conducted using an online survey between January and July 2021 during the second and third waves of the coronavirus pandemic in South Africa. The online survey collected data on mental health symptoms, experiences of stress and social support, COVID-19 infection history, perceptions of COVID-19, and household conditions. The survey was administered in English and distributed using online listservs, social media, community groups, and non-profit organizations. Organizations working with resource-constrained communities were targeted to increase representation of the sample and reduce selection bias, given the online nature of the survey. Inclusion criteria were as follows: adults 18 years of age and older; English proficiency; lived in South Africa for at least three weeks during the pandemic; and ability to provide informed consent. Participants provided formal written consent. Individual participants were not identifiable during or after the survey data collection process. All study procedures were approved by the University of the Witwatersrand Human Research Ethics Committee.

### Study measures

Participants first completed surveys querying demographic and household information. Socioeconomic status was assessed using an asset inventory of the following household items: cell phone, computer, electricity, internet access, landline telephone, microwave, motor vehicle, pay television, radio, refrigerator, television, video machine, and washing machine. Education was assessed by querying participants to report the highest level of schooling completed.

COVID-19 psychosocial stress was assessed using an ethnographically derived survey tool based on in-depth ethnographic interviews with 55 adults in the metropolitan Johannesburg region, 12 adults living in rural Thohoyandou in Limpopo Province, and participant observation for eight months during the COVID-19 pandemic in Johannesburg (Kim *in prep*). Interviews and field notes were thematically analyzed and twenty of the most prevalent and salient stressors were identified and converted into items for the COVID-19 psychosocial stress scale. Items included stressors related to health (feeling unsafe, having a chronic or existing health condition), socioeconomic adversity (unemployment, food insecurity, financial insecurity), socialization (not being able to socialize, not being able to attend gatherings), and resource deficits (lack of transportation, difficulty accessing healthcare), among others. Participants reported the degree to which each item served as a source of stress based on a 4-point Likert scale, which included the following responses: “Never, Seldom, Sometimes, Often, Very Often.” The Likert scale for each item ranged from 0-4. All items were summed to create a total score of COVID-19 psychosocial stress. The internal reliability for this scale was acceptable (α = 0.79).

Mental health outcomes were assessed using four Likert scale-based surveys, which assessed symptoms of four separate psychiatric conditions: depression, anxiety, post-traumatic stress disorder (PTSD), and bipolar disorder. The internal reliability for all measures passed the threshold for acceptability (α > 0.7). Depressive symptoms were assessed using the Patient Health Questionnaire (PHQ-9), a nine-item survey that measures common symptoms of depression, such as fatigue, irritability, melancholia, and trouble concentrating (α = 0.93). Anxiety symptoms were measured using the General Anxiety Disorder Scale (GAD-7), a seven-item survey that assesses key symptoms of anxiety, including nervousness, rumination, and restlessness, among others (α = 0.94).

PTSD symptoms were assessed using the PTSD Checklist – Civilian Version, which comprises 17 questions that query key disease symptoms (α = 0.96). While PTSD diagnoses typically query symptoms in response to a particular event, the PCL-C assesses PTSD symptoms related to a set of “stressful experiences” experienced by the individual and can be viewed as a screening tool for PTSD symptoms. Finally, bipolar disorder symptoms were assessed using the Mood Disorder Questionnaire (MDQ), a screening tool for bipolar symptoms, including increased energy, grandiosity, decreased need for sleep, and others. The first thirteen items of the MDQ were summed to create a composite score of bipolar disorder symptomatology (α = 0.86). The following cut-off scores were used for the following measures: ≥10 (Patient Health Questionnaire-9), ≥10 (Generalised Anxiety Disorder-7), ≥31 (PTSD Checklist - Civilian Version), and ≥7 (Mood Disorder Questionnaire)

Social support was evaluated using the Multidimensional Scale for Perceived Social Support (MSPSS). The MSPSS is a 12-item tool that measures perceptions of support from family, friends, and significant others (Zimet et al., 1988). Finally, coping resources were assessed using an ethnographically derived coping measure (developed through the same procedure described above), which assessed the availability and use of a variety of psychological skills, social practices, economic resources, and other tools utilized to cope with the pandemic.

### Statistical analysis

Data were analyzed using Stata 15.1 (College Station, TX). Bivariate associations were conducted to estimate the relationships between COVID-19 psychosocial stress, all adult mental health measures, social support, coping, and covariates. We then fitted linear regression models to the data and ran four separate sets of analyses based on the specified mental health outcomes: depression, anxiety, PTSD, and bipolar disorder symptoms. COVID-19 psychosocial stress was the primary exposure variable of interest, and social support and coping resources were treated as moderators of the association between COVID-19 psychosocial stress and adult mental health. Psychological, household, and social factors that were thought to potentially confound the relationship between COVID-19 psychosocial stress and adult mental health were included as covariates: age, gender, assets, education, adverse childhood experiences, exercise, disease status, and hours worked. Individuals missing relevant data needed for this analysis were excluded through listwise deletion.

## RESULTS

Full data were available for n=117 individuals out of a total of n=395 who were eligible for the study, provided informed consent, participated in data collection. Table 1 describes the characteristics of our analytic sample. The average age was 36.8 years, 83% of the sample was female, and a majority of the sample had some post-secondary education. The average number of adverse childhood experiences was 2.4, and the average COVID-19 psychosocial stress score was 22.7 (out of 80). The average score for depressive symptoms was 9.3 (PHQ-9), 8.6 for anxiety symptoms (GAD-9), 39.1 for PTSD symptoms (PCL-C), and 3.8 for bipolar symptoms (MDQ). The prevalence rates of probable psychiatric disorders across the following psychopathologies in our sample are as follows: 38% for depression, 39% for anxiety, 57% for PTSD, and 21% for bipolar disorder.

**Table 1.** Demographic characteristics, social experience, and mental health

| Variables | n = 117 | % | Range |
| --- | --- | --- | --- |
| *Demographics* |  |  |  |
| Age | 36.8 (11.7) |  | 21-76 |
| Gender |  |  |  |
| Male | 23 | 19.7 |  |
| Female | 91 | 77.8 |  |
| Non-binary/Genderqueer | 3 | 2.5 |  |
| Education |  |  |  |
| Finished high school & matric | 9 | 7.7 |  |
| Some university | 15 | 12.8 |  |
| Graduated university | 93 | 79.5 |  |
| Assets | 10.5 (2.0) |  | 1-13 |
| *Social experience & mental health* |  |  |  |
| COVID-19 psychosocial stress | 22.7 (11.4) |  | 1-55 |
| Adverse childhood experiences | 2.4 (2.1) |  | 0-9 |
| Perceived social support | 61.0 (16.2) |  | 15-84 |
| Coping resources | 28.2 (7.9) |  | 6-49 |
| Exercise (hours) | 8.8 (11.2) |  | 0-60 |
| Average hours of work per week | 34.7 (16.5) |  | 0-72 |
| Number of chronic conditions | 0.35 (0.7) |  | 0-3 |
| Depressive symptoms | 9.3 (7.3) |  | 0-27 |
| Depression caseness ($\geq 10$) | 45 | 38.5 | |
| Anxiety symptoms | 8.7 (6.5) |  | 0-21 |
| Anxiety caseness ($\geq 10$) | 46 | 39.3 | |
| PTSD symptoms | 39.1 (18.3) |  | 17-85 |
| PTSD caseness ($\geq 30$) | 82 | 57.3 | |
| Bipolar disease symptoms | 3.8 (3.5) |  | 0-13 |
| Bipolar caseness ($\geq 7$) | 24 | 20.5 | |

Supplementary Tables 1 and 2 report forms of social support received from friends, family, and significant others and the availability of coping resources. The most prevalent forms of social support included the presence of “a special person with whom I can share my joys and sorrows,” “a special person who is a real source of comfort to me,” “a special person in my life who cares about my feelings,” “a special person who is around when I am in need,” and “friends with whom I can share my joys and sorrows”. The most common forms of coping were receiving support from family, staying occupied/busy, sleeping, receiving support from friends, and having a positive outlook.

All bivariate analyses between COVID-19 psychosocial stress and all four mental health outcomes were positive (depression: b = 0.30; anxiety: b = 0.23; PTSD = 0.81; bipolar disorder: b = 0.12) and highly significant *p*<0.0001. Fully adjusted models found that COVID-19 stress remained directly associated with all four mental health outcomes, and all associations were significant (*p* < 0.01) (depression: b = 0.21; anxiety: b = 0.17; PTSD: b = 0.54; bipolar symptoms: b = 0.074) (not shown). Identification as female was positively associated with worse depression, anxiety, and PTSD scores (*p* < 0.05). Older age was associated with lower anxiety scores (b = −0.10; *p* = 0.49) while adverse childhood experiences (b = 2.4; *p* = 0.001) and the number of chronic diseases (b = 5.7; *p* = 0.021) were associated with higher PTSD scores. Older age (b = −0.065; *p* = 0.022) and educational attainment (b = −1.03; *p* = 0.024) were associated with lower bipolar disorder scores and the number of hours worked was associated with higher bipolar disorder (b = 0.052; *p* = 0.01).

Table 2a shows the moderating effect of social support on the association between COVID-19 psychosocial stress and mental health. After adjusting for covariates, social support did not significantly buffer the mental health effects of COVID-19 psychosocial stress (depression: b = -.0010, *p* = 0.75; anxiety: b = -.000073, *p* = 0.98; PTSD: b = −0.0084, *p* = 0.28; bipolar disorder: b = −0.0022; *p* = 0.20). The R^2^ for each of the models is as follows: 35% for depression, 32% for anxiety, 47% for PTSD, and 32% for bipolar disorder.

**Table 2a.** Regression models predicting buffering role of social support on mental health impacts of COVID-19 psychosocial stress

|  | Depressive symptoms (PHQ-9) |  |  | Anxiety symptoms (GAD-7) |  |  |
| --- | --- | --- | --- | --- | --- | --- |
|  | b | SE | 95% CI | b | SE | 95% CI |
| COVID-19 psychosocial stress | 0.24 | 0.22 | -0.19, 0.67 | 0.15 | 0.20 | -0.24, 0.54 |
| Social support | -0.078 | 0.090 | -0.26, 0.099 | -0.080 | 0.081 | -0.24, 0.082 |
| COVID-19 stress x social support | -0.0011 | 0.0034 | -0.0079, 0.0057 | -0.000073 | 0.0031 | -0.0063, 0.0061 |
| Age (years) | -0.087 | 0.057 | -0.20, 0.027 | -0.10* | 0.052 | -0.21, -0.0013 |
| Female | 2.58 | 1.36 | -0.11, 5.27 | 2.41 | 1.23 | -0.038, 4.85 |
| Education | -1.15 | 0.93 | -2.99, 0.70 | -0.59 | 0.84 | -2.26, 1.09 |
| Assets | 0.079 | 0.31 | -0.53, 0.69 | 0.13 | 0.28 | -0.42, 0.68 |
| ACEs | 0.30 | 0.31 | -0.32, 0.91 | 0.32 | 0.28 | -0.24, 0.88 |
| Chronic disease | 1.30 | 1.07 | -0.83, 3.42 | 1.41 | 0.97 | -0.52, 3.35 |
| Exercise | 0.024 | 0.055 | -0.086, 0.13 | -0.034 | 0.050 | -0.13, -0.066 |
| Hours worked | -0.019 | 0.042 | -0.10, 0.06 | -0.0083 | 0.038 | -0.084, 0.067 |
| Constant | 19.7 | 11.6 | -3.39, 42.8 | 15.2 | 10.6 | -5.76, 36.1 |

*Note:* ACEs = Adverse childhood experiences; *b* = unstandardized regression weights; CI = confidence interval; \* $p < 0.05$ ; \*\* $p < 0.01$ ; \*\*\* $p < 0.001$

|  | PTSD symptoms (PCL-C) |  |  | Bipolar symptoms (MDQ) |  |  |
| --- | --- | --- | --- | --- | --- | --- |
|  | b | SE | 95% CI | b | SE | 95% CI |
| COVID-19 psychosocial stress | 0.99* | 0.48 | 0.027, 1.95 | 0.20 | 0.11 | -0.0096, 0.41 |
| Social support | 0.024 | 0.20 | -0.37, 0.42 | 0.033 | 0.044 | -0.054, 0.12 |
| COVID-19 stress x social support | -0.0084 | 0.0077 | -0.024, 0.0069 | -0.0022 | 0.0017 | -0.0055, 0.0012 |
| Age (years) | -0.21 | 0.13 | -0.47, 0.43 | -0.063* | 0.028 | -0.12, -0.0070 |
| Female | 6.7* | 3.05 | 0.66, 12.7 | 0.94 | 0.67 | -0.38, 2.26 |
| Education | -1.6 | 2.1 | -5.75, 2.53 | -0.90 | 0.46 | -1.8, 0.0067 |
| Assets | 0.44 | 0.69 | -0.92, 1.8 | -0.049 | 0.15 | -0.35, 0.25 |
| ACEs | 2.3** | 0.69 | 0.89, 3.6 | 0.22 | 0.15 | -0.078, 0.52 |
| Chronic disease | 5.2* | 2.4 | 0.42, 10.0 | 0.89 | 0.53 | -0.15, 1.9 |
| Exercise | 0.13 | 0.12 | -0.12, 0.37 | 0.046 | 0.027 | -0.0079, 0.010 |
| Hours worked | 0.016 | 0.094 | -0.17, 0.20 | 0.046* | 0.021 | 0.0050, 0.087 |
| Constant | 29.3 | 26.1 | -22.5, 81.0 | 7.2 | 5.7 | -4.1, 18.6 |

*Note:* ACEs = Adverse childhood experiences; *b* = unstandardized regression weights; CI = confidence interval; \* $p < 0.05$ ; \*\* $p < 0.01$ ; \*\*\* $p < 0.001$

Table 2b reports the moderating effect of coping resources on the relationship between COVID-19 psychosocial stress and mental health. In fully adjusted models, coping significantly buffered against symptoms of depression (b = −0.014, *p* = 0.043; see fig 1), anxiety (b = −0.013, *p* = 0.038; see fig 2), and PTSD (b = −0.030, *p* = 0.044; see fig 3), but not bipolar symptoms (b = - 0.0045, *p* = 0.18). The R^2^ for each of the models is as follows: 37% for depression, 34% for anxiety, 51% for PTSD, and 33% for bipolar disorder.

**Figure 1.**
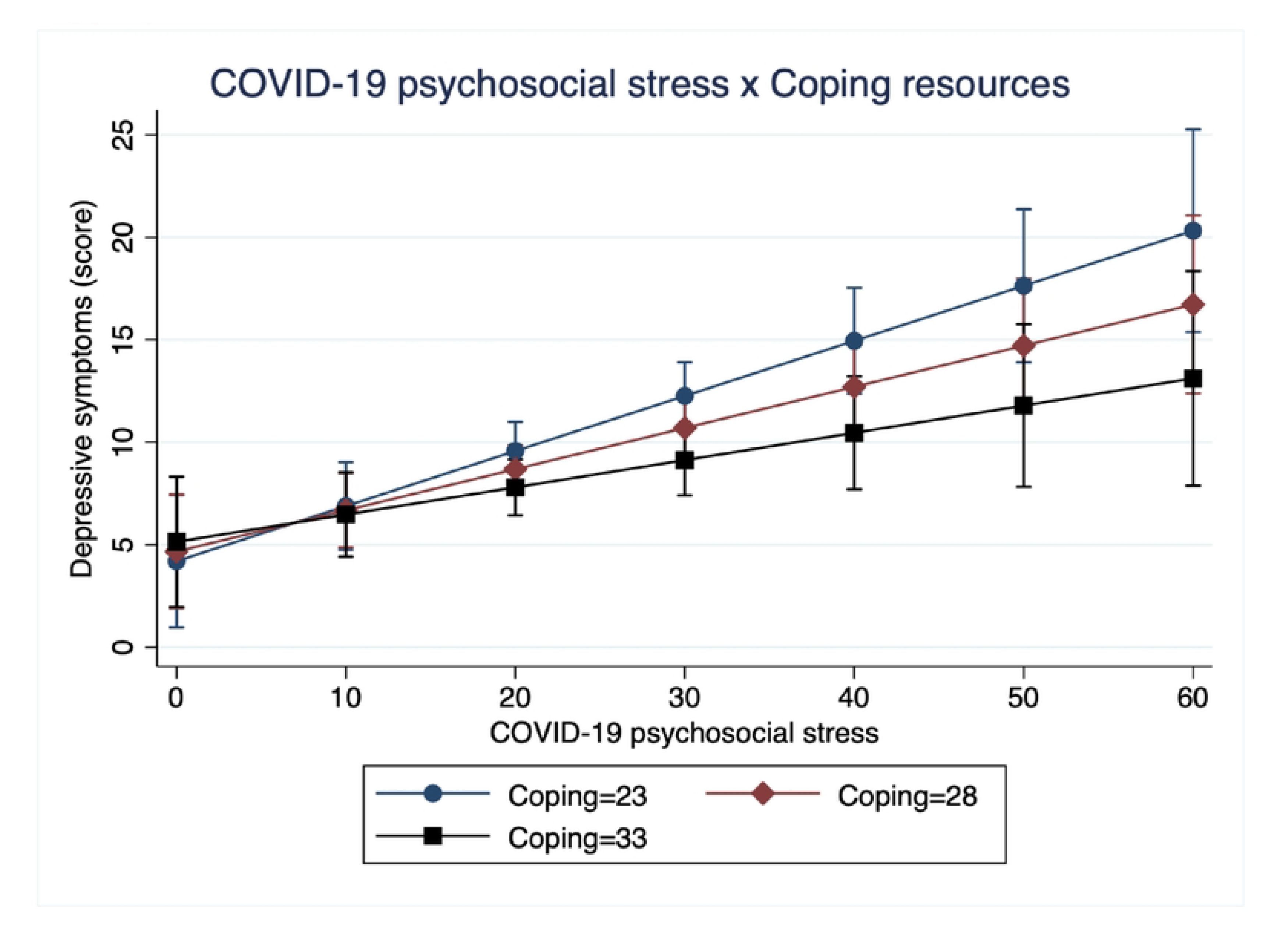
Interaction between COVID-19 psychosocial stress and coping resources predicting depressive symptoms.

**Figure 2.**
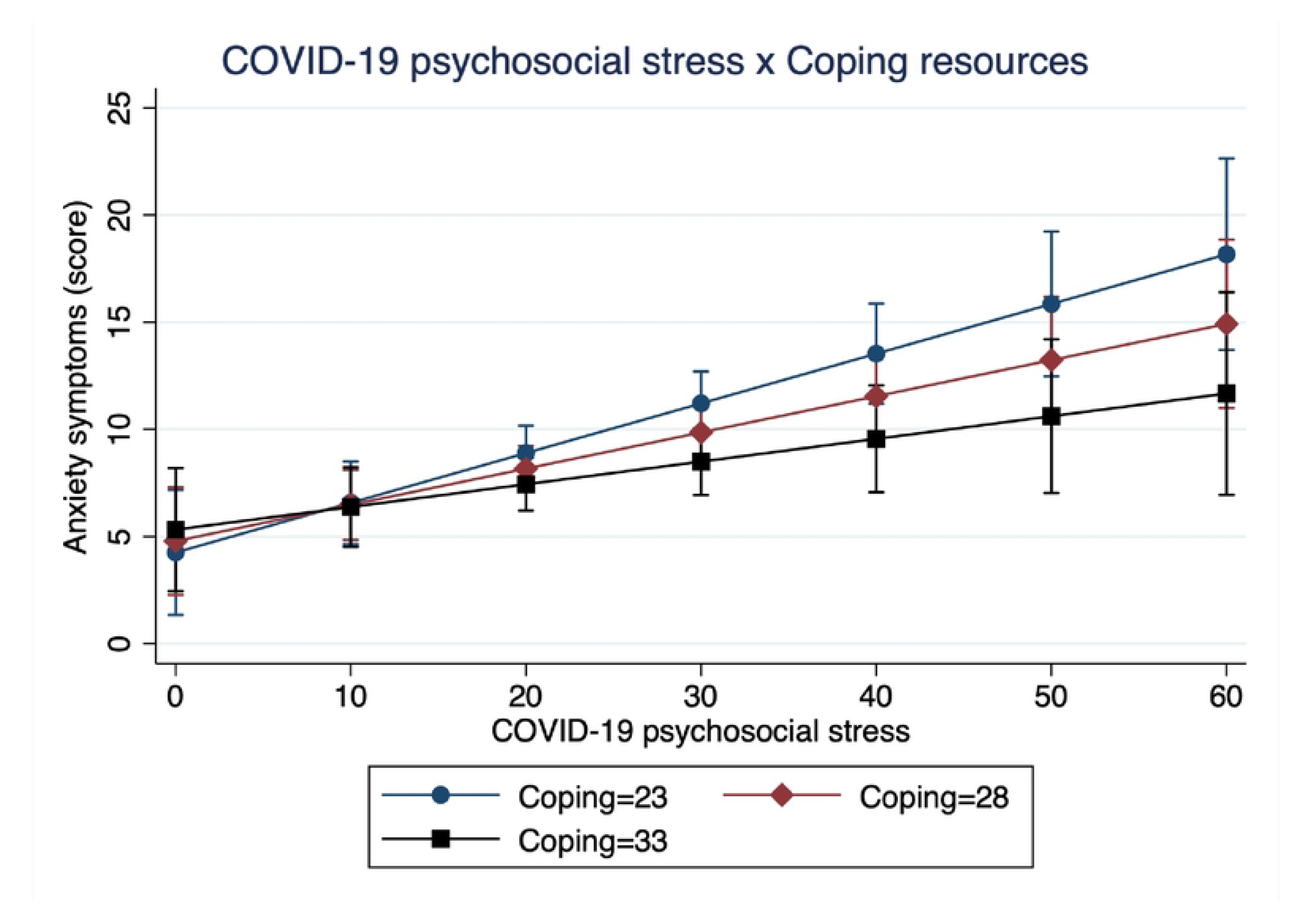
Interaction between COVID-19 psychosocial stress and coping resources predicting depressive symptoms.

**Figure 3.**
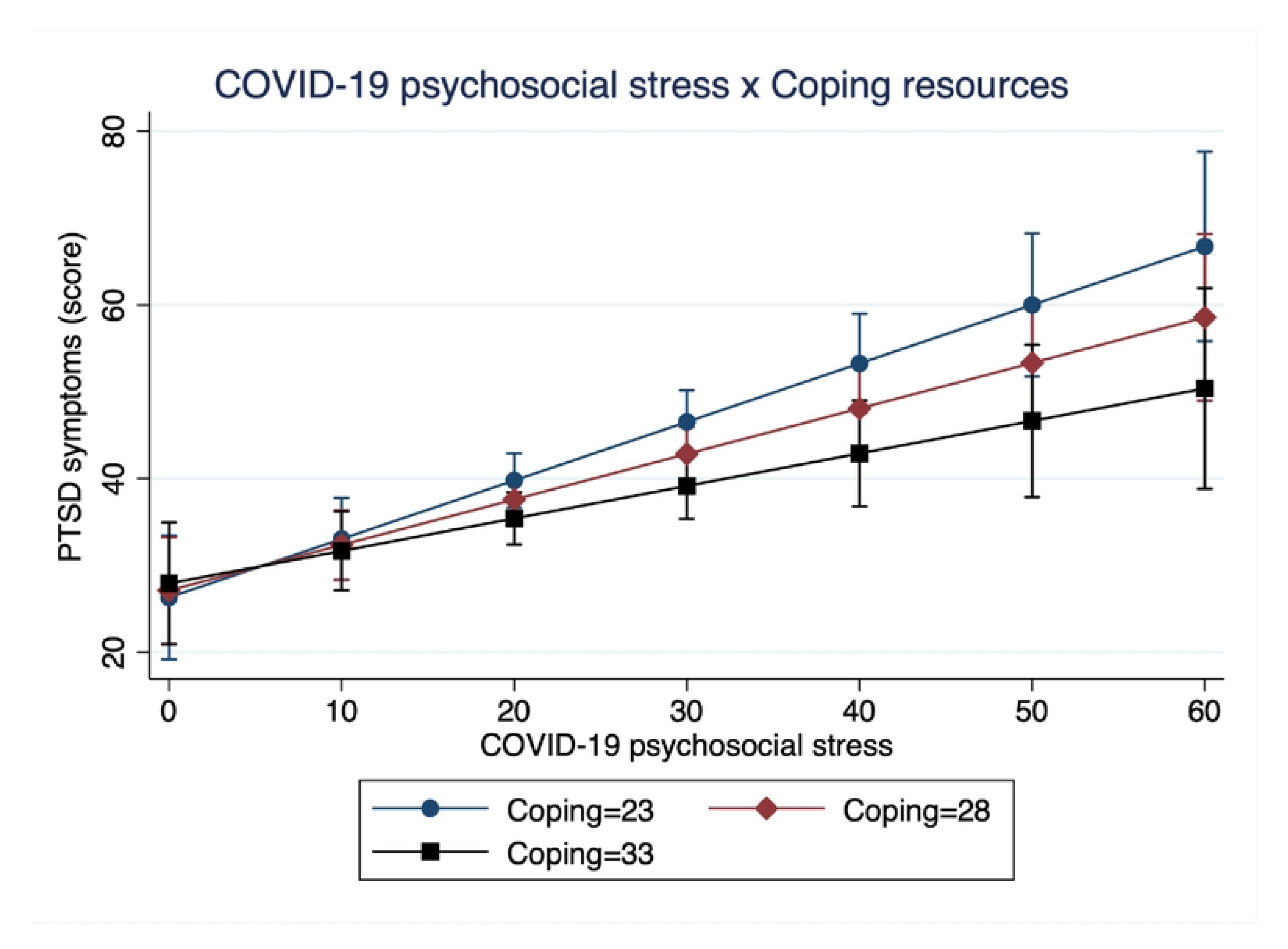
Interaction between COVID-19 psychosocial stress and coping resources predicting depressive symptoms.

**Table 2b.** Regression models predicting buffering role of coping resources on mental health impacts of COVID-19 psychosocial stress

|  | Depressive symptoms (PHQ-9) |  |  | Anxiety symptoms (GAD-7) |  |  |
| --- | --- | --- | --- | --- | --- | --- |
|  | b | SE | 95% CI | b | SE | 95% CI |
| COVID-19 psychosocial stress | 0.58** | 0.19 | 0.20, 0.96 | 0.52** | 0.17 | 0.18, 0.87 |
| Coping resources | 0.095 | 0.16 | -0.22, 0.41 | 0.11 | 0.14 | -0.18, 0.39 |
| COVID-19 stress x coping resources | -0.014* | 0.0066 | -0.027, -0.00043 | -0.013 | 0.0060* | -0.024, -0.00072 |
| Age (years) | -0.073 | 0.056 | -0.19, 0.039 | -0.092 | 0.051 | -0.19, 0.0097 |
| Female | 2.5 | 1.4 | -0.21, 5.2 | 2.3 | 1.2 | -0.14, 4.7 |
| Education | -1.3 | 0.90 | -3.1, 0.46 | -0.68 | 0.82 | -2.3, 0.93 |
| Assets | 0.14 | 0.30 | -0.45, 0.73 | 0.21 | 0.27 | -0.33, 0.74 |
| ACEs | 0.38 | 0.30 | -0.22, 0.98 | 0.39 | 0.27 | -0.15, 0.93 |
| Chronic disease | 1.1 | 1.1 | -0.97, 3.2 | 1.3 | 0.96 | -0.65, 3.2 |
| Exercise | -0.020 | 0.058 | -0.14, 0.095 | -0.075 | 0.053 | -0.18, 0.029 |
| Hours worked | -0.0015 | 0.040 | -0.081, 0.078 | 0.0030 | 0.036 | -0.069, 0.075 |
| Constant | 11.8 | 10.4 | -8.9, 32.5 | 6.3 | 9.4 | -12.4, 25.0 |

*Note:* ACEs = Adverse childhood experiences; *b* = unstandardized regression weights; CI = confidence interval; \* $p < 0.05$ ; \*\* $p < 0.01$ ; \*\*\* $p < 0.001$

|  | PTSD symptoms (PCL-C) |  |  | Bipolar symptoms (MDQ) |  |  |
| --- | --- | --- | --- | --- | --- | --- |
|  | b | SE | 95% CI | b | SE | 95% CI |
| COVID-19 psychosocial stress | 1.4** | 0.42 | 0.52, 2.2 | 0.20* | 0.095 | 0.0093, 0.39 |
| Coping resources | 0.16 | 0.35 | -0.54, 0.86 | 0.038 | 0.079 | -0.12, 0.20 |
| COVID-19 stress x coping resources | -0.030* | 0.015 | -0.059, -0.00089 | -0.0045 | 0.0033 | -0.011, 0.0021 |
| Age (years) | -0.19 | 0.12 | -0.43, 0.01 | -0.060* | 0.028 | -0.12, -0.0051 |
| Female | 6.2* | 3.0 | 0.23, 12.1 | 0.79 | 0.67 | -0.54, 2.1 |
| Education | -2.0 | 2.0 | -6.0, 1.9 | -0.97* | 0.45 | -1.9, -0.085 |
| Assets | 0.61 | 0.66 | -0.70, 1.9 | -0.037 | 0.15 | -0.33, 0.26 |
| ACEs | 2.4** | 0.67 | 1.1, 3.7 | 0.23 | 0.15 | -0.062, 0.53 |
| Chronic disease | 4.8* | 2.3 | 0.12, 9.4 | 0.87 | 0.53 | -0.18, 1.9 |
| Exercise | 0.020 | 0.13 | -0.23, 0.28 | 0.031 | 0.29 | -0.027, 0.088 |
| Hours worked | 0.051 | 0.089 | -0.13, 0.23 | 0.051* | 0.020 | 0.011, 0.090 |
| Constant | 26.3 | 23.1 | -19.4, 72.1 | 8.7 | 5.2 | -1.60, 18.9 |

*Note:* ACEs = Adverse childhood experiences; *b* = unstandardized regression weights; CI = confidence interval; \* $p < 0.05$ ; \*\* $p < 0.01$ ; \*\*\* $p < 0.001$

## DISCUSSION

In this analysis of adults living under the COVID-19 pandemic in South Africa, coping resources buffered against the poor mental health effects of COVID-19 psychosocial stress, whereas perceived social support did not significantly moderate the association between COVID-19 stress and adult mental health. Specifically, greater use of coping resources attenuated symptoms of depression, anxiety, and PTSD due to COVID-19 stress. We also found elevated levels of poor mental health in this sample during the COVID-19 pandemic and lockdown. We highlight the fact that this sample is overly educated, largely female, and represents a group of adults with moderate to high socioeconomic status. Despite the difficult and prolonged conditions of the pandemic, our results suggest that adults effectively utilized resources to positively cope with the various stressors brought on by the pandemic.

The buffering role of coping resources is consistent with the larger literature on adult mental health during the pandemic and various other conditions of psychosocial stress. Specifically, our results build on past studies that show that coping behaviors buffer against symptoms of adult depression, anxiety, and PTSD due to psychosocial stress from the pandemic (Okafor et al. 2021; Senger 2023; Suhail et al. 2022). Past studies have shown that a variety of coping resources is valuable, including cognitive strategies (e.g., positive thinking, reappraisal), behavioral practices, (e.g., handwashing, information gathering), social capital (e.g., structural, instrumental), and spirituality (e.g., praying, connectedness, meaning-making) (Pankowski & Wytrychiewicz-Pankowska 2023). This important set of mental health-sparing behaviors protected numerous communities at high risk of COVID-19 infection and those with pre-existing vulnerabilities, including frontline healthcare workers, adults living with chronic diseases, and elderly populations (Hong et al. 2023; Lábadi et al. 2022; Tahara et al. 2021). Coping also provided similar buffering effects against psychosocial stress among adults in past pandemics, including Ebola (James et al. 2019) and HIV/AIDS (Seffren et al. 2018).

These results also add to the growing literature in South Africa that report the positive mental health benefits of coping during the COVID-19 lockdown (Eloff, 2021; Engelbrecht et al., 2021; Kim et al. 2022; Paredes-Ruvalcaba et al. 2023; Scheunemann et al. 2023; van der Merwe et al., 2021; Visser & Law-van Wyk, 2021). While a majority of these studies focus on healthcare workers or university students, rather than community-based adults such as those included in our sample, the buffering effect of coping resources is consistent across analyses. Engelbrecht et al. (2021) found that preparedness for care for COVID-19 patients, avoidance-based coping, and current health status before COVID-19 predicted lower PTSD symptoms in nurses deployed during the pandemic. In a qualitative study of community-based adults in Gauteng, Paredes-Ruvalcaba et al. (2023) found that adults from diverse racial and socioeconomic groups utilized a variety of coping strategies to overcome the stressors of the pandemic, such as peer support, prayer, exercise, financial support, mindset reframing, natural remedies, and following COVID-19 protocols. Notably, coping resources may have positive impacts on mental health but poor, longer-term effects on physical health. For instance, Visser & Law-van Wyk (2021) reported that South African university students engaged in substance use to cope with the pandemic, despite early social policies prohibiting the sale of cigarettes and alcohol in the country. Together, these studies and our results suggest that South Africans utilized a variety of coping mechanisms to protect themselves against the negative mental health impacts of COVID-19 psychosocial stress.

We also found that perceived social support did not significantly buffer against the adverse psychological effects of the pandemic, which contradicts the larger literature that highlights the positive mental health effects of adult social support (Harandi et al. 2017; Kessler & McLeod 1985). Studies worldwide have repeatedly shown the protective and buffering effects of social support against a wide range of poor mental health outcomes, including depression, anxiety, and suicidal ideation (Casale et al. 2015; Chang et al. 2017; Olashore et al. 2021). Additionally, studies in South Africa have described the importance of receiving social support from family, friends, significant others, and coworkers. Paredes-Ruvalcaba et al. (2023) found that South African adults utilized various means of virtual communication, such as video calls, group texts, social media, and online services, to provide emotional support, process negative cognitions, and stay hopeful. Scheunemann et al. (2023) described the vital role of social relationships and active coordination between psychiatric healthcare workers in Gauteng to provide more tangible, instrumental support among one another, including the organization of online prayer groups, alternative work schedules to cover missing shifts due to pandemic-related health problems and pooled financial resources.

Despite these past findings, we find that social support did not buffer against the poor mental health effects of COVID-19 psychosocial stress. Given the state-enforced isolation and disruption of infrastructural support systems, drastic changes in social behaviors and structures during the pandemic may have altered the role of social support on health in this setting and may explain these null findings. Past studies have shown that co-occurring emotional and psychological experiences, such as feelings of loneliness, negative mood, and problematic social relationships, can compromise the positive mental health effects of social support (Wang et al. 2018). For instance, conditions of depression and anxiety can negatively bias an individual’s appraisal of their social relationships, leading to altered evaluations of their interpersonal contexts. Additionally, the Multidimensional Scale of Perceived Social Support may not fully capture the social and interpersonal dynamics between the respondent and the relationships in question (e.g., family, friends, and significant others). Past research has described many familial and social relationships shifted during the lockdown, and greater strain in social relationships, both familial and non-familial, predicted worse mental health outcomes during the pandemic (Essler et al. 2021; Randall et al. 2021; Skinner et al. 2021).

### Limitations

Our study is not without limitations. Our findings are not generalizable to the entire South African population as our sample represents a relatively wealthy, educated, and majority female set of adults. The online nature of data collection likely biased our sample to those who had access to the internet, computers, and other socioeconomic resources, leading to a privileged sample. The cross-sectional design of our analysis may also subject our analysis to reverse causality, limiting our ability to determine the true temporal ordering of events. Finally, different forms of social support important for buffering against the effects of stress may not have been captured by our social support measure.

## CONCLUSION

In this online study of 117 adults during the second and third waves of the COVID-19 pandemic in South Africa, we found that the use of coping resources, but not perceived social support alone, significantly buffered against worse symptoms of depression, anxiety, and post-traumatic stress disorder. This sample of South African adults exhibited elevated levels of mental health symptoms, with more than half of the sample reporting PTSD symptoms, over a third exhibiting symptoms of depression and anxiety, and a fifth of adults reporting symptoms of bipolar disorder. These data suggest that adults utilized a variety of coping resources to protect their mental health against psychosocial stress experienced during the COVID-19 lockdown and pandemic in South Africa.

## Data Availability

We are unable to share pertinent data at the time of editorial and peer review, but we plan to make the data that support the findings of this study available on request from the corresponding author. The data are not publicly available due to privacy or ethical restrictions.

## Data Availability Statement

The data that support the findings of this study are available on request from the corresponding author. The data are not publicly available due to privacy or ethical restrictions.

## BIBLIOGRAPHY

Asante, K. O. Social support and the psychological wellbeing of people living with HIV/AIDS in Ghana. African Journal of Psychiatry (South Africa). 2012; 15(5), 340–345. https://doi.org/10.4314/ajpsy.v15i5.42

Barbarin, O. A., & Richter, L. M. Mandela’s children: Growing up in post-apartheid South Africa. Routledge; 2013

Bhattacharjee, A., & Ghosh, T. COVID-19 Pandemic and Stress: Coping with the New Normal. Journal of Prevention and Health Promotion. 2022; 3(1), 30–52. https://doi.org/10.1177/26320770211050058

Brakefield, W. S., Olusanya, O. A., White, B., & Shaban-Nejad, A. Social Determinants and Indicators of COVID-19 among Marginalized Communities: A Scientific Review and Call to Action for Pandemic Response and Recovery. Disaster Medicine and Public Health Preparedness. 2022; 617, 9–11. https://doi.org/10.1017/dmp.2022.104

Braveman, P., & Gottlieb, L. The social determinants of health: It’s time to consider the causes of the causes. Public Health Reports. 2014; 129(SUPPL. 2), 19–31. https://doi.org/10.1177/00333549141291s206

Brener, L., Broady, T., Cama, E., Hopwood, M., de Wit, J. B., & Treloar, C. The role of social support in moderating the relationship between HIV centrality, internalised stigma and psychological distress for people living with HIV. AIDS care. 2020; 32(7), 850–857.

Burke, H. M., Davis, M. C., Otte, C., & Mohr, D. C. Depression and cortisol responses to psychological stress: A meta-analysis. Psychoneuroendocrinology. 2005; 30(9), 846–856. https://doi.org/10.1016/j.psyneuen.2005.02.010

Cacioppo, J. T., Cacioppo, S., Capitanio, J. P., & Cole, S. W. The Neuroendocrinology of Social Isolation. Annual Review of Psychology. 2015; 66(1), 733–767. https://doi.org/10.1146/annurev-psych-010814-015240

Casale, M., Wild, L., Cluver, L., & Kuo, C. Social support as a protective factor for depression among women caring for children in HIV-endemic South Africa. Journal of behavioral medicine. 2015; 38, 17–27.

Chang, Q., Chan, C. H., & Yip, P. S. A meta-analytic review on social relationships and suicidal ideation among older adults. Social science & medicine. 2017; 191, 65–76.

Charney, D. S. Psychobiological Mechanisms of Resilience and Vulnerability: Implications for Successful Adaptation to Extreme Stress. American Journal of Psychiatry. 2004; 161, 195–216. https://doi.org/10.1176/appi.ajp.161.2.195

Chew, Q. H., Wei, K. C., Vasoo, S., & Sim, K. Psychological and Coping Responses of Health Care Workers Toward Emerging Infectious Disease Outbreaks: A Rapid Review and Practical Implications for the COVID-19 Pandemic. Singapore Med J. [revista en Internet] 2020 [acceso 10 de marzo de 2021]. 2020; 61(7): 3. Singapore Med J, 61(7), 350–356.

Cohen, S., & Wills, T. A. Stress, Social Support, and the Buffering Hypothesis. Psychological Bulletin. 1985; 98(2), 310–357. https://doi.org/10.1016/0163-8343(94)90083-3

Coovadia, H., Jewkes, R., Barron, P., Sanders, D., & McIntyre, D. The health and health system of South Africa: historical roots of current public health challenges. The Lancet. 2009; 374(9692), 817–834. https://doi.org/10.1016/S0140-6736(09)60951-X

Eloff, I. College students’ well-being during the COVID-19 pandemic: An exploratory study. Journal of Psychology in Africa. 2021; 31(3), 254–260.

Essler, S., Christner, N., & Paulus, M. Longitudinal relations between parental strain, parent– child relationship quality, and child well-being during the unfolding COVID-19 pandemic. Child Psychiatry & Human Development. 2021; 52(6), 995–1011.

Fang, X., Li, S., Yu, H., Wang, P., Zhang, Y., Chen, Z., … Ma, X. Epidemiological, comorbidity factors with severity and prognosis. Aging. 2020; 12(13), 12493–12503.

Fino, E., Bonfrate, I., Fino, V., Bocus, P., Russo, P. M., & Mazzetti, M. Harnessing distress to boost growth in frontline healthcare workers during COVID-19 pandemic: the protective role of resilience, emotion regulation and social support. Psychological Medicine. 2023; 53(2), 600–602.

Fu, M., Guo, J., & Zhang, Q. The associations of pandemic-related difficulties with depressive symptoms and psychological growth among American older adults: Social support as moderators. Journal of health psychology. 2022; 13591053221124374.

Guilaran, J., de Terte, I., Kaniasty, K., & Stephens, C. Psychological Outcomes in Disaster Responders: A Systematic Review and Meta-Analysis on the Effect of Social Support.

International Journal of Disaster Risk Science. 2018; 9(3), 344–358. https://doi.org/10.1007/s13753-018-0184-7

Harandi, T. F., Taghinasab, M. M., & Nayeri, T. D. The correlation of social support with mental health: A meta-analysis. Electronic physician. 2017; 9(9), 5212.

Hennessy, M. B., Kaiser, S., & Sachser, N. Social buffering of the stress response: Diversity, mechanisms, and functions. Frontiers in Neuroendocrinology. 2009; 30(4), 470–482. https://doi.org/10.1016/j.yfrne.2009.06.001\

Hong, C., Queiroz, A., & Hoskin, J. The impact of the COVID-19 pandemic on mental health, associated factors and coping strategies in people living with HIV: a scoping review. Journal of the International AIDS Society. 2023; 26(3), e26060.

Hossain, M. M., Tasnim, S., Sultana, A., Faizah, F., Mazumder, H., Zou, L., … Ma, P. Epidemiology of mental health problems in COVID-19: A review. F1000Research. 2020; 9, 1–16. https://doi.org/10.12688/f1000research.24457.1

Hostinar, C. E., Sullivan, R. M., & Gunnar, M. R. Psychobiological Mechanisms Underlying the Social Buffering of the HPA Axis: A Review of Animal Models and Human Studies across Development. Psychological Bulletin. 2015; 140(1), 1–47. https://doi.org/10.1037/a0032671.Psychobiological

Hou, T., Zhang, T., Cai, W., Song, X., Chen, A., Deng, G., & Ni, C. Social support and mental health among health care workers during Coronavirus Disease 2019 outbreak: A moderated mediation model. Plos one. 2020; 15(5), e0233831.

James, P. B., Wardle, J., Steel, A., & Adams, J. Post-Ebola psychosocial experiences and coping mechanisms among Ebola survivors: a systematic review. Tropical Medicine & International Health. 2019; 24(6), 671–691.

Jones, D. L., Ballivian, J., Rodriguez, V. J., Uribe, C., Cecchini, D., Salazar, A. S., … & Alcaide, M. L. Mental health, coping, and social support among people living with HIV in the Americas: a comparative study between Argentina and the USA during the SARS-CoV-2 pandemic. AIDS and Behavior. 2021; 25, 2391–2399.

Kaminer, D., & Eagle, G. Traumatic stress in South Africa. Wits University Press.

Kessler, R. C., & McLeod, J. D. (1985). Social support and mental health in community samples. 2010; Academic Press.

Kim, A. W., Said Mohamed, R., Norris, S. A., Richter, L. M., & Kuzawa, C. W. Psychological legacies of intergenerational trauma under South African apartheid: Prenatal stress predicts greater vulnerability to the psychological impacts of future stress exposure during late adolescence and early adulthood in Soweto, South Africa. Journal of Child Psychology and Psychiatry and Allied Disciplines. 2023; 64(1), 110–124. https://doi.org/10.1111/jcpp.13672

Kim, A.W., Maaroganye, K., & Subramaney, U. Mental health experiences of public psychiatric healthcare workers during COVID-19 across southern Gauteng, South Africa: a call for strengthening. South African Health Review. 2021; 2021(1), 143-151.

Kim, A. W., Nyengerai, T., & Mendenhall, E. Evaluating the mental health impacts of the COVID-19 pandemic: Perceived risk of COVID-19 infection and childhood trauma predict adult depressive symptoms in urban South Africa. Psychological medicine. 2022; 52(8), 1587–1599.

Kokou-Kpolou, C. K., Derivois, D., Rousseau, C., Balayulu-Makila, O., Hajizadeh, S., Birangui, J. P., … & Cénat, J. M. Enacted Ebola Stigma and Health-related Quality of Life in Post Ebola Epidemic: A Psychosocial Mediation Framework Through Social Support, Self-Efficacy, and Coping. Applied Research in Quality of Life. 2022; 17(5), 2809–2832.

Lábadi, B., Arató, N., Budai, T., Inhóf, O., Stecina, D. T., Sík, A., & Zsidó, A. N. Psychological well-being and coping strategies of elderly people during the COVID-19 pandemic in Hungary. Aging & Mental Health. 2022; 26(3), 570–577.

Labrague, L. J. Psychological resilience, coping behaviours and social support among health care workers during the COVID-19 pandemic: A systematic review of quantitative studies. Journal of nursing management. 2021; 29(7), 1893–1905.

Liu, C., Huang, N., Fu, M., Zhang, H., Feng, X. L., & Guo, J. Relationship between risk perception, social support, and mental health among general Chinese population during the COVID-19 pandemic. Risk management and healthcare policy. 2021; 1843–1853.

Lupien, S J, McEwen, B. S., Gunnar, M. R., & Heim, C. Effects of stress throughout the lifespan on the brain, behaviour and cognition. Nature Reviews Neuroscience. 2009; 10(6), 434–445. https://doi.org/10.1038/nrn2639

Lupien, Sonia J., Juster, R. P., Raymond, C., & Marin, M. F. The effects of chronic stress on the human brain: From neurotoxicity, to vulnerability, to opportunity. Frontiers in Neuroendocrinology. 2018; 49(February), 91–105. https://doi.org/10.1016/j.yfrne.2018.02.001

Manchia, M., Gathier, A. W., Yapici-Eser, H., Schmidt, M. V., de Quervain, D., van Amelsvoort, T., … Vinkers, C. H. The impact of the prolonged COVID-19 pandemic on stress resilience and mental health: A critical review across waves. European Neuropsychopharmacology. 2022; 55, 22–83. https://doi.org/10.1016/j.euroneuro.2021.10.864

Marin, M. F., Lord, C., Andrews, J., Juster, R. P., Sindi, S., Arsenault-Lapierre, G., … Lupien, S. J. Chronic stress, cognitive functioning and mental health. Neurobiology of Learning and Memory. 2011; 96(4), 583–595. https://doi.org/10.1016/j.nlm.2011.02.016

McEwen, B. S. Neurobiological and Systemic Effects of Chronic Stress. Chronic Stress. 2017; 1. https://doi.org/10.1177/2470547017692328

Meewisse, M. L., Reitsma, J. B., De Vries, G. J., Gersons, B. P. R., & Olff, M. Cortisol and post-traumatic stress disorder in adults: Systematic review and meta-analysis. British Journal of Psychiatry. 2007; 191(NOV.), 387–392. https://doi.org/10.1192/bjp.bp.106.024877

Metzger, L. J., Carson, M. A., Lasko, N. B., Paulus, L. A., Orr, S. P., Pitman, R. K., & Yehuda, R. Basal and suppressed salivary cortisol in female Vietnam nurse veterans with and without PTSD. Psychiatry Research. 2008; 161(3), 330–335. https://doi.org/10.1016/j.psychres.2008.04.020

Okafor, C. N., Bautista, K. J., Asare, M., & Opara, I. Coping in the time of COVID-19: Buffering stressors with coping strategies. Journal of Loss and Trauma. 2022; 27(1), 83–91.

Olashore, A. A., Akanni, O. O., & Oderinde, K. O. Neuroticism, resilience, and social support: correlates of severe anxiety among hospital workers during the COVID-19 pandemic in Nigeria and Botswana. BMC Health Services Research. 2021; 21, 1–7.

Our World in Data. COVID-19 Data explorer. 2023; https://ourworldindata.org/explorers/coronavirus-data-explorer

Oyenubi, A., Kim, A. W., & Kollamparambil, U. COVID-19 risk perceptions and depressive symptoms in South Africa: Causal evidence in a longitudinal and nationally representative sample. Journal of Affective Disorders. 2022; 308, 616–622.

Pankowski, D., & Wytrychiewicz-Pankowska, K. Turning to religion during COVID-19 (Part I): A systematic review, meta-analysis and meta-regression of studies on the relationship between religious coping and mental health throughout COVID-19. 2023; Journal of religion and health, 1–34.

Polizzi, C., Lynn, S. J., & Perry, A. Stress and coping in the time of COVID-19: Pathways to resilience and recovery. Clinical Neuropsychiatry. 2020; 17(2), 59–62. https://doi.org/10.36131/CN20200204

Randall, A. K., Leon, G., Basili, E., Martos, T., Boiger, M., Baldi, M., … & Chiarolanza, C. Coping with global uncertainty: Perceptions of COVID-19 psychological distress, relationship quality, and dyadic coping for romantic partners across 27 countries. Journal of Social and Personal Relationships. 2022; 39(1), 3–33.

Russell, G., & Lightman, S. The human stress response. Nature Reviews Endocrinology. 2019; 15(9), 525–534. https://doi.org/10.1038/s41574-019-0228-0

Sanford, L. D., Wellman, L. L., Adkins, A. M., Guo, M.-L., Zhang, Y., Ren, R., … Tang, X. Modeling integrated stress, sleep, fear and neuroimmune responses: Relevance for understanding trauma and stress-related disorders. Neurobiology of Stress. 2023; 23(June 2022), 100517. https://doi.org/10.1016/j.ynstr.2023.100517

Sanyaolu, A., Okorie, C., Marinkovic, A., & Patidar, R. Comorbidity and its impact on patients with COVID-19. SN Comprehensive Clinical Medicine. 2020; 2, 1069–1076.

Schierberl Scherr, A. E., Ayotte, B. J., & Kellogg, M. B. Moderating roles of resilience and social support on psychiatric and practice outcomes in nurses working during the COVID-19 pandemic. SAGE open nursing. 2021; 7, 23779608211024213.

Seffren, V., Familiar, I., Murray, S. M., Augustinavicius, J., Boivin, M. J., Nakasujja, N., … & Bass, J. Association between coping strategies, social support, and depression and anxiety symptoms among rural Ugandan women living with HIV/AIDS. AIDS care. 2018; 30(7), 888–895.

Senger, A. R. Hope’s Relationship with Resilience and Mental Health During the COVID-19 Pandemic. Current Opinion in Psychology. 2023; 101559.

Skinner, A. T., Godwin, J., Alampay, L. P., Lansford, J. E., Bacchini, D., Bornstein, M. H., … & Yotanyamaneewong, S. Parent–adolescent relationship quality as a moderator of links between COVID-19 disruption and reported changes in mothers’ and young adults’ adjustment in five countries. Developmental Psychology. 2021; 57(10), 1648.

Steudte, S., Kirschbaum, C., Gao, W., Alexander, N., Schönfeld, S., Hoyer, J., & Stalder, T. Hair cortisol as a biomarker of traumatization in healthy individuals and posttraumatic stress disorder patients. Biological Psychiatry. 2013; 74(9), 639–646. https://doi.org/10.1016/j.biopsych.2013.03.011

Stiegler, N., & Bouchard, J. P. South Africa: Challenges and successes of the COVID-19 lockdown. Annales Medico-Psychologiques. 2020; 178(7), 695–698. https://doi.org/10.1016/j.amp.2020.05.006

Subramaney, U., Kim, A. W., Chetty, I., Chetty, S., Jayrajh, P., Govender, M., … & Pak, E. Coronavirus disease 2019 (COVID-19) and psychiatric sequelae in South Africa: Anxiety and beyond. Wits journal of clinical medicine. 2020; 2(2), 115–122.

Suhail, A., Dar, K. A., & Iqbal, N. COVID-19 related fear and mental health in Indian sample: The buffering effect of support system. Current Psychology. 2022; 41(1), 480–491.

Szkody, E., Stearns, M., Stanhope, L., & McKinney, C. Stress-Buffering Role of Social Support during COVID-19. Family Process. 2021; 60(3), 1002–1015. https://doi.org/10.1111/famp.12618

Tahara, M., Mashizume, Y., & Takahashi, K. Coping mechanisms: exploring strategies utilized by Japanese healthcare workers to reduce stress and improve mental health during the COVID- 19 pandemic. International journal of environmental research and public health. 2021; 18(1), 131.

Taylor, S. E. Social support: A review. In H. S. Friedman (Ed.), Oxford handbook of health psychology. 2011; (pp. 192–217). New York, NY: Oxford University Press.

Uchino, B. N. Social Support and Health: A Review of Physiological Processes Potentially Underlying Links to Disease Outcomes. Journal of Behavioral Medicine. 2006; 29(4). https://doi.org/10.1007/s10865-006-9056-5

Uchino, B. N., Cacioppo, J. T., & Kiecolt-glaser, J. K. The Relationship Between Social Support and Physiological Processes: A Review with Emphasis on Underlying Mechanisms and Implications for Health. Psychological Bulletin. 1996; 119(3), 488–531. https://doi.org/10.1097/JGP.0b013e3181f7d89a

van der Merwe, L., Erasmus, E., Morelli, J., Potgieter, H., Modise, J., Viviers, E., … & Lerumo, K. Stories of the role musicking plays in coping with the COVID-19 pandemic in South Africa. Psychology of Music. 2021; 03057356211026959.

Visser, M., & Law-van Wyk, E. University students’ mental health and emotional wellbeing during the COVID-19 pandemic and ensuing lockdown. South African Journal of Psychology. 2021; 51(2), 229–243.

Wang, B., Li, R., Lu, Z., & Huang, Y. Does comorbidity increase the risk of patients with COVID-19. Aging. 2020; 12(7), 6049–6057.

Wang, J., Mann, F., Lloyd-Evans, B., Ma, R., & Johnson, S. Associations between loneliness and perceived social support and outcomes of mental health problems: a systematic review. BMC psychiatry. 2018; 18(1), 1–16.

Yehuda, R., Golier, J. A., & Kaufman, S. Circadian rhythm of salivary cortisol in Holocaust survivors with and without PTSD. American Journal of Psychiatry. 2005; 162(5), 998–1000. https://doi.org/10.1176/appi.ajp.162.5.998

Zaken, M. D., Boyraz, G., & Dickerson, S. S. COVID-19 pandemic-related stressors and posttraumatic stress: The main, moderating, indirect, and mediating effects of social support. Stress and Health. 2022; 38(3), 522–533.

